# Infectious Complications in Philadelphia-Negative Adolescent and Young Adult Acute Lymphoblastic Leukemia Patients in the Era of Pediatric-Inspired Regimen

**DOI:** 10.1101/2025.01.23.25321023

**Authors:** Phoutthasin Vongngakesone, Piangrawee Niprapan, Sirichai Srichairatanakool, Nonthakorn Hantrakun, Teerachat Punnachet, Sasinee Hantrakool, Pokpong Piriyakhuntorn, Chatree Chai-Adisaksopha, Ekarat Rattarittamrong, Adisak Tantiworawit, Lalita Norasetthada, Thanawat Rattanathammethee

**Author notes:** Corresponding author (TR).

## Abstract

**Introduction:** Pediatric acute lymphoblastic leukemia (ALL) has cure rates exceeding 90%, while adult ALL has a 3-year survival rate of only 40–50%. Adolescents and young adults (AYAs) face unique challenges due to age-related factors and high-risk disease features. Although pediatric-inspired regimens improve survival, intensive chemotherapy increases susceptibility to severe infections.

**Methods:** A retrospective analysis was conducted on AYA-ALL patients treated at Chiang Mai University Hospital from 1 January 2007 to 31 December 2023. Patients were classified into pediatric-inspired (TPOG) or adult regimens (Hyper-CVAD or GMALL). Infections were categorized as clinically documented infections (CDI) or microbiologically documented infections (MDI). Multivariate logistic regression was used to identify infection risk factors.

**Results:** Among 94 patients (62.8% male; median age 22.9 years), 56.4% received TPOG and 43.6% received adult regimens. Infectious complications occurred in 79.8% of patients, with higher rates in the adult regimen group (90.2% vs. 71.7%, *p* = 0.03). CDI was more frequent in the adult group (73.4% vs. 52.8%, *p* = 0.04), while fungal infections were more common in the TPOG group (26.4% vs. 9.8%, *p* = 0.04). Adult regimens significantly increased infection risk (OR 3.55; 95% CI 1.02–12.36, *p* = 0.04). Infection rates peaked during induction (47.8%) and consolidation (51.8%). During induction, MDI was lower in the adult group (21% vs. 79%, *p* = 0.01), while CDI was higher during consolidation (69% vs. 30%, *p* < 0.01). Gram-negative bacteria were the most common pathogens (85%), particularly *E. coli* (27%) and *Salmonella* spp. (21%). Fungal infections affected 19.1% of patients, with invasive pulmonary aspergillosis being the most common (11.7%).

**Conclusion:** AYA-ALL patients are highly susceptible to infections, especially those receiving adult regimens. Fungal infections were notably more common in patients on the TPOG regimen. Strengthening infection prevention and providing early treatment are vital for improving patient outcomes.

## Introduction

Acute lymphoblastic leukemia (ALL) is the most common pediatric cancer but is relatively rare in adults, with an incidence of 1.7 cases per 100,000 annually in the total U.S. population [1]. Over the past few decades, the prognosis for pediatric ALL has significantly improved, with cure rates exceeding 90% using standard intensive multi-agent chemotherapy protocols [2, 3]. However, adult ALL remains associated with poorer outcomes, with only 40-50% survival at three years [4]. This survival gap is likely due to differences in disease biology, treatment response, and patient-related factors.

Adolescents and young adults (AYAs), aged 15 to 39 years, face distinct treatment challenges due to their transitional physiology and a higher prevalence of high-risk disease features, resulting in event-free survival rates of 30-45% [5-7]. The presence of specific genetic factors, diversity within this age range, and different treatment protocols may contribute to the lower survival rates observed in adult ALL patients.

Multiple retrospective and prospective studies have demonstrated that pediatric-inspired regimens significantly improve survival outcomes in AYAs compared to standard adult regimens [8-10]. Despite improved survival, infections remain a significant cause of morbidity and mortality, particularly during induction therapy, affecting over 30% of patients. Bacterial infections accounted for nearly 70% of infection-related deaths, while fungal infections accounted for about 20% [11].

In Thailand, a study involving 107 ALL patients (67.3% received adult regimens and 32.7% received pediatric-inspired regimens) found that patients treated with pediatric-inspired protocols had better two-year disease-free survival (47.1% vs. 24.7%, hazard ratio [HR] 1.73, 95% CI 1.22–3.03) and overall survival (50.8% vs. 31.2%, HR 1.52, 95% CI 0.83–2.78) compared to those treated with adult regimens [12]. Similarly, a multi-centre prospective study showed that AYAs had significantly higher two-year event-free survival compared to adults with ALL (45.5% vs 26.2%; P < 0.019) [13].

Despite evidence supporting pediatric-inspired regimens, the infection risks and associated factors in AYAs with ALL remain understudied, particularly when comparing pediatric-inspired and adult regimens in Thailand.

## Methods

### Study design and Patients

This retrospective cohort study included newly diagnosed adolescent and young adult patients with acute lymphoblastic leukemia (AYA-ALL), aged 15-40 years, treated at Chiang Mai University Hospital between 1 January 2007 and 31 December 2023. The diagnosis was confirmed according to the 2016 WHO Classification of Tumors of Hematopoietic and Lymphoid Tissues [14]. This study followed the Declaration of Helsinki and the International Conference on Harmonization Guidelines for Good Clinical Practice. The institutional ethical review board of the Faculty of Medicine, Chiang Mai University, Thailand, approved the study (study code: MED-2566-0630). Informed consent was waived due to the retrospective design and anonymized data collection.

Patients received induction chemotherapy using either a pediatric-inspired regimen (a modified Thai Pediatric Oncology Group [TPOG] protocol) [12, 15], a dosage and intensity-adjusted version of the Thai national childhood ALL protocol, or adult regimens (Hyper-CVAD or GMALL) [16, 17]. Patients were excluded if they did not receive high-intensity chemotherapy, had Philadelphia chromosome-positive ALL and/or the *BCR::ABL1* fusion gene (detected via polymerase chain reaction), had incomplete clinical records, received induction therapy prior to referral, or had a known HIV infection. All patients received standard antimicrobial prophylaxis with trimethoprim/sulfamethoxazole for *Pneumocystis jirovecii* pneumonia (PJP) and acyclovir for herpes simplex virus (HSV) infections. Due to limited availability of mold-active antifungal agents, fluconazole was used for fungal prophylaxis. However, fluconazole and trimethoprim/sulfamethoxazole were withheld during certain treatment phases to prevent drug interactions. Granulocyte colony-stimulating factor (G-CSF) was routinely administered during induction and consolidation phases to reduce neutropenia-related infection risk. Notably, specific data on antimicrobial prophylaxis regimens for bacterial, fungal, and viral infections were not analyzed in this study.

### Data collection and outcomes

Clinical data were retrospectively collected from electronic medical records with de-identified patients’ data, potential bias from missing data. Data included baseline demographics, laboratory results, infectious events, and treatment outcomes. Infectious complications during all chemotherapy phases (induction, consolidation, and maintenance) were analyzed.

The primary outcome was the prevalence of infectious complications among AYA-ALL patients. The secondary outcomes were to identify associated risk factors for infectious complications and compare the infectious complications rate between patients receiving pediatric-inspired (TPOG) and adult regimens (Hyper-CVAD or GMALL).

### Definitions of infectious complications

Infectious complications were categorized as follows:

- **Clinically Documented Infection (CDI):** Clinical signs and symptoms of infection without an identifiable pathogen.
- **Microbiologically Documented Infection (MDI):** Isolation of a pathogen from blood cultures or other infection sites.

Fever was defined as a body temperature >38.3°C or a temperature >38°C persisting for at least 1 hour. Febrile neutropenia was defined as fever with an absolute neutrophil count (ANC) < 500/µL [18].

### Sample size and statistical analysis

Based on previous studies [9, 10], an infection prevalence of 50% was assumed in AYA-ALL patients. The required sample size was 97 patients, with a margin of error of 10% [19]. Continuous variables were reported as mean with standard deviation (SD) or median with interquartile range (IQR), depending on data distribution. Categorical variables were expressed as counts and percentages. Group comparisons for categorical data were performed using the chi-square test or Fisher’s exact test. Continuous variables were compared using the Student’s *t*-test or Mann-Whitney *U* test, as appropriate. Variables with a p-value < 0.1 in univariate analysis were entered into a backward stepwise multivariate logistic regression model to identify independent risk factors for infections. Results were reported as odds ratios (OR) and 95% confidence intervals (CI). Statistical significance was set at p-value < 0.05. Patients with missing data were excluded. All analyzes were performed using Microsoft Excel 2013 and STATA Version 17 (StataCorp, TX, USA) software.

## Results

### Demographic and clinical characteristics

A total of 124 newly diagnosed Philadelphia-negative ALL patients at Chiang Mai University Hospital between 1 January 2007 and 31 December 2023 were retrospectively reviewed, 30 patients were excluded, and 94 AYA-ALL patients were recruited **(Fig 1)**. The majority were male (62.8%, n = 59), with a median age of 22.9 years (IQR 15-39.8). B-ALL was the most common subtype (58.8%). Of these, 56.4% (n = 53) received pediatric-inspired TPOG regimen, while 43.6% (n = 41) received adult regimens (24 on Hyper-CVAD and 17 on GMALL). Overall, the median follow-up was 14.3 months (IQR 7.5-30.9). Relapse or refractory disease occurred in 55.3% of patients, with a significantly higher rate in the adult regimen group compared to the TPOG group (68.3 % vs. 45.3, *p* = 0.02). Only four patients (4.3%) underwent allogeneic stem cell transplantation (AlloSCT). Baseline characteristics were summarized in **Table 1**.

**Fig 1.**
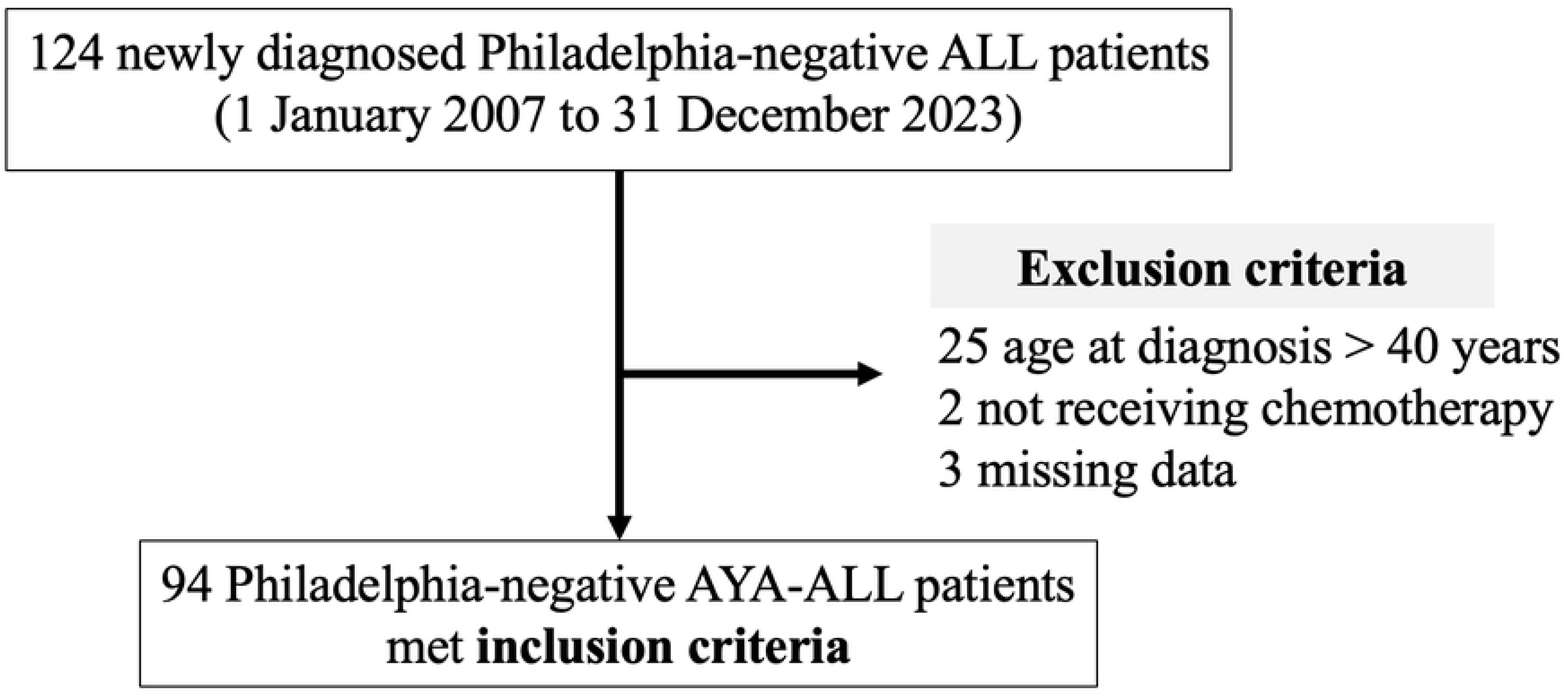
Flow diagram demonstrated the recruitment alongside of inclusion and exclusion criteria.

**Table 1.** Demographic and clinical characteristics of AYA-ALL patients.

| Variables | All patients<br>(n = 94) | TPOG<br>(n = 53) | Adult regimen<br>(n = 41) | *p*-value |
| --- | --- | --- | --- | --- |
| Age, years | 22.9 (15-39.8) | 23.8(15-39.2) | 21.7 (15-39.8) | 0.63 |
| Male, n (%) | 59 (62.8) | 36 (67.9) | 23 (56.1) | 0.24 |
| ALL subtype, n (%) |  |  |  | 0.31 |
| B-ALL | 55 (58.5) | 31 (58.5) | 24 (58.5) |  |
| T-ALL | 37 (39.4) | 22 (41.5) | 15 (36.6) |  |
| N/A | 2 (2.1) | 0 | 2 (4.9) |  |
| Hemoglobin, g/dL | 9.5 (7.5-11.4) | 9.5 (7.4-11.7) | 9.1 (7.5-11.2) | 0.60 |
| WBC, /μL | 16600<br>(4500-47500) | 23000<br>(5200-51400) | 14170<br>(3570-39400) | 0.40 |
| Platelet, /μL | 75900<br>(32000-145000) | 84000<br>(33000-158000) | 65000<br>(32000-139000) | 0.73 |
| Peripheral blood blasts, % | 49.5 (4-83) | 48 (4.7-85) | 50 (3-82) | 0.82 |
| Bone marrow blasts, % | 90 (80-90) | 90(80-90) | 90(80-90) | 0.12 |
| Induction death, n (%) | 4 (4.2) | 2 (3.8) | 2 (4.8) | 1 |
| Proceed to consolidation phase, n (%) | 81 (86.1) | 44 (83.0) | 37 (90.2) | 0.31 |
| Duration of consolidation, days | 223 (93-313) | 261.5 (109.5- 398.5) | 199 (66-245) | <0.01 |
| Proceed to maintenance phase, n (%) | 31 (32.9) | 18 (33.9) | 13 (31.7) | 0.81 |
| Duration of maintenance, days | 721 (301-770) | 731 (385-862) | 516 (147-744) | 0.16 |
| Completed maintenance phase, n (%) | 16 (51.6) | 10 (55.5) | 6 (46.1) | 0.72 |
| Relapse/Refractory, n (%) | 52 (55.3) | 24 (45.3) | 28 (68.3) | 0.02 |
| Allogeneic SCT, n (%) | 4 (4.3) | 3 (5.7) | 1 (2.4) | 0.62 |

As the continuous data in this study did not follow a normal distribution, all data were reported as medians with interquartile ranges (IQR). WBC: white blood cell count, SCT: stem cell transplantation

### Infectious outcomes

Infectious complications were reported in 79.8% (n = 75) of patients. Clinically documented infections (CDI) occurred in 61.7% (n = 58), and microbiologically documented infections (MDI) in 55.3% (n = 52). Bacterial infections were the most common (44.6%, n = 42), followed by fungal infections (19.1%, n = 18).

Patients on adult regimens experienced significantly more infectious complications than those on the TPOG regimen (90.2% vs. 71.7%, *p* = 0.03), with higher CDI rates (73.4% vs. 52.8%, *p* = 0.04). Conversely, fungal infections were more frequent in the TPOG group (26.4% vs. 9.8%, *p* = 0.04) **(Table 2).**

**Table 2.** Infectious complications among AYA-ALL patients based on treatment regimens.

| Variables | All patients<br>(n=94) | TPOG<br>(n=53) | Adult regimen<br>(n=41) | *p*-value |
| --- | --- | --- | --- | --- |
| Overall infection, n (%) | 75 (79.8) | 38 (71.7) | 37 (90.2) | 0.03 |
| CDI, n (%) | 58 (61.7) | 28 (52.8) | 30 (73.2) | 0.04 |
| MDI, n (%) | 52 (55.3) | 28 (52.8) | 24 (58.5) | 0.58 |
| Bacterial infection, n (%) | 42 (44.6) | 22 (41.5) | 20 (48.8) | 0.48 |
| Fungus infection, n (%) | 18 (19.1) | 14 (26.4) | 4 (9.8) | 0.04 |

CDI: clinically documented infection, MDI: microbiologically documented infection

Infection rates peaked during induction (47.8%) and consolidation (51.8%). During consolidation, infections were significantly higher in the adult regimen group compared to the TPOG group (64.9% vs. 40.9%, *p* = 0.03) **(Fig 2)**. MDI was most frequent in the TPOG group during induction (79% vs. 21%, *p* = 0.01), whereas CDI was more common in the adult regimen group during the consolidation (69% vs. 30%, *p* < 0.01) (**S1 Table)**. Details of CDI based on treatment group and phase of treatment were provided **(S2 Table).**

**Fig 2.**
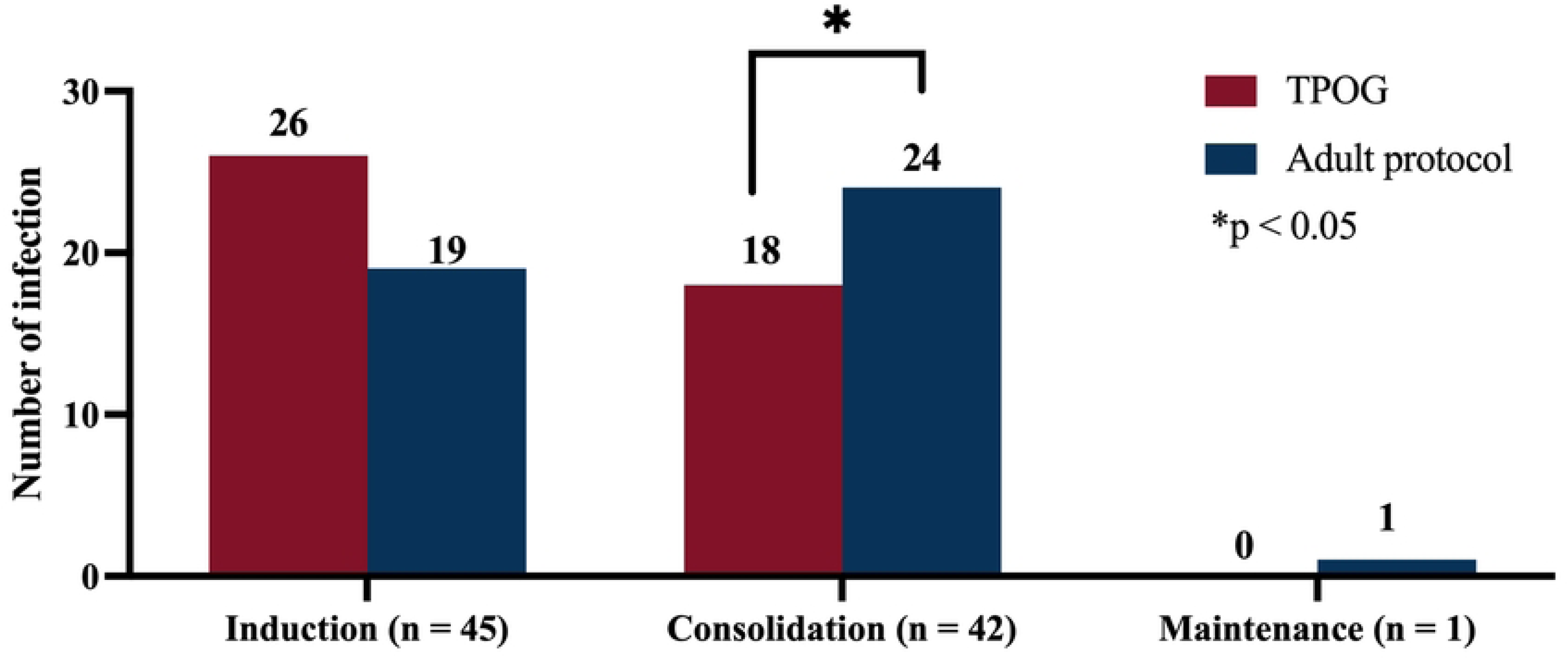
Number of infection based on treatment regimens and treatment phases.

The most common infection sites for MDI were the bloodstream (40%), lung (24%) and the genitourinary tract (16%) **(S3 Table)**. Bacterial infections were the most prevalent (73%), followed by fungal (24%) and viral (3%) infections. Gram-negative bacteria dominated (85%) among bacterial pathogens, with *E. coli* (27%), *Salmonella spp.* (21%), and *Klebsiella spp.* (19%) being the most prevalent. *Aspergillus spp.* was the most common fungal pathogen **(S4 Table and S5 Table).** Induction-related mortality was 4.2% (n = 4), with septicemia causing death in 2 patients and hemorrhagic complications in 2 patients.

### Risk factors for infectious complications

Multivariate logistic regression identified treatment with an adult regimen as a significant risk factor for infectious complications among AYA-ALL patients (OR 3.55, 95% CI 1.02–12.36, *p* = 0.04) **(Table 3)**. For MDI, female sex (OR 3.88, 95% CI 1.41–10.68, *p* < 0.01) and platelet counts below 75,000 /μL (OR 3.41, 95% CI 1.33–8.74, *p* = 0.01) were independently associated with a higher risk. In contrast, patients younger than 23 years (OR 0.33, 95% CI 0.12–0.85, *p* = 0.02) and those receiving only first-line treatment (OR 0.36, 95% CI 0.13–0.93, *p* = 0.03) had a significantly lower risk of MDI **(S6 Table)**.

**Table 3.** Univariate and multivariate analyses of risk factor for infectious complications.

| Variables |  | Univariate |  |  | Multivariate |  |  |
| --- | --- | --- | --- | --- | --- | --- | --- |
| | | Infection<br>(n=75) | Non infection<br>(n=19) | $p$-value | Odds<br>ratio | 95% CI | $p$-value |
| Treatment regimens | Adults | 37 (90.2) | 4 (9.8) | 0.026 | 3.55 | 1.02-12.36 | 0.04 |
|  | TPOG | 38 (71.7) | 15 (28.3) |  |  |  |  |
| Sex | Female | 30 (85.7) | 5 (14.3) | 0.2 |  |  |  |
|  | male | 45 (76.3) | 14 (23.7) |  |  |  |  |
| ALL type | B cell | 44 (80) | 8 (20.5) | 0.95 |  |  |  |
|  | others | 31 (79.5) | 11 (20.0) |  |  |  |  |
| Relapse | yes | 44 (84.6) | 8 (15.4) | 0.19 |  |  |  |
|  | no | 31 (73.8) | 11 (26.2) |  |  |  |  |
| 1<sup>st</sup> line of treatment | yes | 39 (72.2) | 15 (27.8) | 0.03 | 0.4 | 0.11-1.40 | 0.155 |
|  | no | 36 (90) | 4 (10) |  |  |  |  |
| Age, years | < 23 | 37 (78.7) | 10 (21.3) | 0.79 |  |  |  |
| | ≥ 23 | 38 (80.8) | 9 (19.1) | | | | |
| WBC, /μL | <16600 | 40 (85.1) | 7 (14.9) | 0.19 | | | |
| | $\geq 16600$ | 35 (74.5) | 12 (25.5) | | | | |
| Hb, g/dL | $< 9.5$ | 38 (80.9) | 9 (19.1) | 0.79 | | | |
| | $\geq 9.5$ | 37 (78.7) | 10 (21.3) | | | | |
| Platelet, /μL | $< 75000$ | 39 (84.8) | 7 (15.2) | 0.23 | | | |
| | $\geq 75000$ | 36 (75.0) | 12 (25.0) | | | | |
| PB blasts, % | $< 50\%$ | 42 (89.4) | 5 (10.6) | 0.02 | 3.41 | 1.05-11.04 | 0.04 |
| | $\geq 50\%$ | 33 (70.2) | 14 (29.8) | | | | |
| BM blasts, % | $< 90\%$ | 34 (79.0) | 9 (20.9) | 0.87 | | | |
| | $\geq 90\%$ | 41 (80.4) | 10 (19.6) | | | | |

WBC: white blood cell count, Hb: hemoglobin, PB: peripheral blood, BM: bone marrow

## Discussion

This study demonstrated a high incidence of infectious complications in AYA-ALL patients, with 79.8% experiencing infections. Clinically documented infections (CDI) occurred in 61.7% of patients, and microbiologically documented infections (MDI) in 55.3%. These rates are comparable to studies from Taiwan and Mexico, which reported infection rates of approximately 87% in ALL patients receiving intensive chemotherapy [20, 21]. Similarly, an Italian study highlighted infectious complications as a leading cause of morbidity in AYA-ALL patients, underscoring the importance of infection prevention strategies [22].

### Phase-specific infection risk

Infection rates were highest during the induction (47.8%) and consolidation (51.8%) phases, aligning with findings from MD Anderson, which reported infection rates of 22% and 63% during these phases, respectively [9]. The increased risk during induction is likely due to the intensive chemotherapy and high-dose steroids, causing severe myelosuppression and immune suppression. During consolidation, cumulative immunosuppression further elevates infection risk. Differences in regimen intensity between TPOG and adult protocols may explain the observed variation in infection rates, highlighting the need for phase-specific supportive care.

### Fungal infection in the TPOG group

Fungal infections were more frequent in patients receiving the TPOG regimen (26.4%) compared to those on adult regimens (9.8%), consistent with findings from Mexico, where 20% of ALL patients developed fungal infections [21]. This increased risk in the TPOG group may result from higher steroid doses and longer chemotherapy durations, which prolong immunosuppression and impair both humoral and cellular immunity. Additionally, fluconazole was used for antifungal prophylaxis due to limited access to mold-active agents at our center, potentially contributing to the higher rates of *Aspergillus* infections. In contrast, studies from Saudi Arabia reported lower fungal infection rates with micafungin or anidulafungin prophylaxis [23]. Environmental factors, particularly in tropical climates, may also increase fungal exposure [24].

### Bacterial infections predominate in adult regimens

Gram-negative bacteria were the most common pathogens (85%), with *E. coli* (27%) and *Salmonella* spp. (21%) being the most prevalent. These findings align with studies from Italy, where gram-negative organisms were predominant in leukemia patients [25]. Patients on adult regimens experienced significantly more bacterial infections, likely due to prolonged and severe neutropenia associated with these protocols.

### Risk factors for infection

Multivariate analysis identified treatment with adult regimens as a significant risk factor for overall infections (OR 3.55; *p*= 0.04). Female gender (OR 3.88; *p* < 0.01) and platelet counts below 75,000/μL (OR 3.41; *p* = 0.01) were independent risk factors for MDI. Conversely, younger patients (<23 years) and those receiving first-line treatment had a lower infection risk. The higher infection rate in females may be due to a higher incidence of urinary tract infections, as reported in other studies [20].

### Clinical implications

These findings emphasize the need for tailored infection prevention strategies based on treatment regimen and phase. For patients receiving pediatric-inspired regimens, particularly TPOG, the use of mold-active antifungal prophylaxis should be considered. Patients on adult regimens may benefit from more aggressive bacterial infection prevention and early intervention during neutropenic episodes. Environmental and regional factors must also be considered when designing infection control protocols.

### Study limitations

This study has several limitations that should be considered when interpreting the findings. Its retrospective design may introduce selection bias, and incomplete or inconsistent medical records could affect data accuracy. The relatively small sample size and single-center setting limit the generalizability of the results to broader populations and other healthcare settings. Additionally, the study did not analyze antimicrobial resistance patterns, which could impact the interpretation of infection outcomes. Variations in antimicrobial prophylaxis regimens for bacterial, fungal, and viral infections were also not fully assessed, potentially influencing infection rates. These limitations highlight the need for larger, multi-center prospective studies to validate these findings and optimize infection prevention strategies in AYA-ALL patients.

## Conclusions

Our study highlights the high burden of infectious complications in AYA-ALL patients, particularly those receiving adult chemotherapy regimens. Fungal infections were notably more common in patients treated with pediatric-inspired regimens, likely due to prolonged steroid use and insufficient mold-active antifungal prophylaxis. These findings underscore the need for targeted infection prevention and early intervention strategies to improve outcomes in this vulnerable population.

## Supporting information

**S1-6 File.** The table S1 to S6 (S1-6_Table.docx)

**S1 Table.** Number and types of infectious episodes (CDI and MDI) based on treatment regimens and treatment phases

**S2 Table.** Clinically documented infection (CDI) based on treatment regimens and treatment phases

**S3 Table.** Sites of microbiologically documented infection (MDI) based on treatment regimens and treatment phases

**S4 Table.** Isolated pathogens of MDI in TPOG regimen group

**S5 Table.** Isolated pathogens of MDI in adult regimen group

**S6 Table.** Univariate and multivariate analyses of risk factor for MDI

## Acknowledgements

I would like to express my sincere gratitude to all the staff at the Division of Hematology, Department of Internal Medicine, Faculty of Medicine, Chiang Mai University, for their invaluable support and dedication to patient care throughout this study.

## Ethics Statement

The study was conducted in accordance with the International Conference on Harmonization for Good Clinical Practice guidelines and the 1964 Declaration of Helsinki. The study was approved by the the institutional ethical review board of the Faculty of Medicine, Chiang Mai University, Thailand (study code: MED-2566-0630). Informed consent was waived due to the retrospective design and anonymized data collection.

## Author’s contributions

Conceptualization: Thanawat Rattanathammethee

Methodology: Thanawat Rattanathammethee

Data curation: Thanawat Rattanathammethee, Phoutthasin Vongngakesone

Formal analysis: Thanawat Rattanathammethee, Phoutthasin Vongngakesone

Investigation: Thanawat Rattanathammethee, Phoutthasin Vongngakesone

Writing-Original draft: Thanawat Rattanathammethee, Phoutthasin Vongngakesone

Writing-review and editing: All authors

## Data Availability Statement

The datasets generated during and/or analyzed in the current study, containing potentially sensitive patient information and de-identified patient data, are available through the research administration section of the Faculty of Medicine at Chiang Mai University, Thailand.

